# Deep Learning Integration of Chest CT Imaging and Gene Expression Identifies Novel Aspects of COPD

**DOI:** 10.1101/2022.09.26.22280242

**Authors:** Junxiang Chen, Xu Zhonghui, Li Sun, Ke Yu, Craig P. Hersh, Adel Boueiz, John Hokanson, Frank C. Sciurba, Edwin K. Silverman, Peter J. Castaldi, Kayhan Batmanghelich

**Author notes:** Correspondence and requests for reprints should be addressed to Junxiang Chen, Department of Biomedical Informatics, University of Pittsburgh, Pittsburgh, 5607 Baum Blvd, Pittsburgh, PA 15206. equal contribution (Peter J. Castaldi, Kayhan Batmanghelich). Author Contributions: The authors meet criteria for authorship as recommended by the International Committee of Medical Journal Editors. J.C., P.J.C, and K.B. designed the study. J.C performed the modeling and statistical analysis and wrote the initial manuscript. Z.X. conducted differential expression and usage analyses. L.S, and K.Y conducted image pre-processing and feature extraction. C.P.H, A.B, J. H., F.C.S, E.K.S and P.J.C assisted with analysis of COPDGene data. All authors contributed to the production of the final manuscript with revision for important intellectual content.

## Abstract

**Rationale:** Chronic obstructive pulmonary disease (COPD) is characterized by pathologic changes in the airways, lung parenchyma, and persistent inflammation, but the links between lung structural changes and patterns of systemic inflammation have not been fully described.

**Objectives:** To identify novel relationships between lung structural changes measured by chest computed tomography (CT) and systemic inflammation measured by blood RNA sequencing.

**Methods:** CT scan images and blood RNA-seq gene expression from 1,223 subjects in the COPDGene study were jointly analyzed using deep learning to identify shared aspects of inflammation and lung structural changes that we refer to as Image-Expression Axes (IEAs). We related IEAs to COPD-related measurements and prospective health outcomes through regression and Cox proportional hazards models and tested them for biological pathway enrichment.

**Measurements and Main Results:** We identified two distinct IEAs: IEA_emph_ captures an emphysema-predominant process with a strong positive correlation to CT emphysema and a negative correlation to FEV_1_ and Body Mass Index (BMI); IEA_airway_ captures an airway-predominant process with a positive correlation to BMI and airway wall thickness and a negative correlation to emphysema. Pathway enrichment analysis identified 29 and 13 pathways significantly associated with IEA_emph_ and IEA_airway_, respectively (adjusted p<0.001).

**Conclusions:** Integration of CT scans and gene expression data identified two IEAs that capture distinct inflammatory processes associated with emphysema and airway-predominant COPD.

**At a Glance Commentary:** *Scientific Knowledge on the Subject:* Chronic obstructive pulmonary disease (COPD) is characterized by lung structural changes and has a prominent systemic inflammatory component, but the links between lung structural changes and patterns of systemic inflammation in COPD have not been fully described.

*What This Study Adds to the Field:* We identified novel relationships between lung structural changes and systemic inflammation by simultaneously analyzing CT scans and blood RNA-sequencing gene expression using deep learning models. We identified two distinct Image-Expression Axes (IEAs) that characterize different inflammatory processes associated with emphysema and airway predominant COPD. This article has an online data supplement, which is accessible from this issue’s table of content online at www.atsjournals.org.

## Introduction

Chronic obstructive pulmonary disease (COPD) is one of the most prevalent chronic diseases (1), responsible for approximately 3 million deaths annually (2). COPD is characterized by persistent respiratory symptoms and poorly reversible airflow limitation (3). It is associated with an abnormal inflammatory response of the lungs to cigarette smoke or other noxious particles (4), which results in lung structural changes, including the loss or narrowing of airways (airway disease) and parenchymal destruction (emphysema) (5). In addition to its characteristic lung structural changes, COPD also has a prominent systemic inflammatory component (6,7).

Although lung structural changes and systemic inflammation are characteristic aspects of COPD, the origin of systemic inflammation and its relation to lung structural changes remain unclear (8). Computed Tomography (CT) scans provide a detailed characterization of lung structural changes (9), and blood gene expression captures systemic inflammatory signals (10). Therefore, we are motivated to understand the relationship between lung structural changes and systemic inflammation by applying a deep learning method to analyze CT imaging and RNA-seq data to identify novel relationships between lung structure changes and systemic inflammation patterns.

Based on the paradigm of COPD as a collection of treatable traits (11), we hypothesize that COPD heterogeneity can be described by continuous measures corresponding to distinct disease processes that are present to varying degrees in affected subjects. We refer to these continuous measures as “disease axes” (12), and we further hypothesize that integrative analysis of CT images and blood RNA-seq data can identify disease axes that identify patterns of association between lung structural abnormalities and systemic inflammation as quantified by blood RNA-seq. We tested these hypotheses by training a deep learning model on data from 1,223 subjects in the COPDGene study (13) with CT scans and blood gene expression data. Our analysis identified two disease axes that capture patterns of CT features consistent with emphysema and airway-predominant disease processes that are also associated with emphysema core-peel distribution and specific inflammatory pathways.

## Methods

A comprehensive description of methods is included in the online supplement, and all analysis code is available in a GitHub repository (https://github.com/batmanlab/IEA).

### Subject Enrollment and Data Collection

COPDGene enrolled 10,198 subjects with a minimum 10 pack-years lifetime smoking history at 21 centers across the United States (NCT00608764, www.copdgene.org) (13). Five-year follow-up data are available for 6,717 subjects, and 10-year follow-up visits are currently being completed. Subjects underwent spirometry, questionnaire assessments, standardized inspiratory and expiratory chest CT imaging, and genome-wide SNP genotyping. At the second visit (year 5), PAXgene blood RNA tubes were collected, and RNA-seq was performed. Each center obtained institutional review board approval, and all participants provided written informed consent.

### Learning Image-Expression Axes (IEAs)

Only subjects whose CT scans were obtained on Siemens scanners with the b31f kernel were analyzed. CT features were extracted from DICOM image files using the following procedure. Each inspiratory chest CT scan was processed into 581 patches with a size of 32^3^ mm^3,^ following the practice in (14). For each patch, 128 features were generated using context-aware self-supervised representation learning (CSRL) (14), producing a matrix of dimension 581 × 128 for each CT scan.

Image-Expression Axes (IEAs) were constructed where CSRL features were the input to a multilayer perceptron (MLP) that produced a low-dimensional representation for each patch. Further supervised dimension reduction was performed to obtain subject-level Image-Expression Axes (IEAs) using a Product of Experts (PoE) model (15). At this stage, we applied statistically independent constraints with the Hilbert-Schmidt independence criterion (HSIC) (16) to ensure that each IEA captured an independent disease process. A final linear layer used IEAs as the input to predict the expression levels of the genes simultaneously. The parameters of the model were jointly optimized via Adam (17). In the training process, we evaluated the impact of feature selection on genes by testing each gene for the association with the top-128 principal components of the CSRL features in the training dataset. We also evaluated various thresholds for gene inclusion determined by the p-value of the F-test for each gene.

We randomly split the data into training and testing sets with sizes of 923 and 300, respectively. Model training was performed in the training set using five-fold cross-validation, giving us five models. The final IEAs were given by taking the average value of the IEAs from the five models.

### Association of IEAs with Clinical Measurements

We computed Pearson correlation coefficients between IEAs and clinical measurements to understand their association. A full description of the measurements is included in the online supplement. Multivariable analyses were conducted by training Ordinary Least Squares (OLS) models for continuous measurements and logistic regression models for categorical measurements. We conducted a survival analysis (starting from the second visit in the supplement)with the Cox proportional hazards model(18). A full description of these models, including model covariates, is provided in the online supplement.

### Comparison of IEAs to Other Disease Axes

We compared IEAs to the following disease axes: 1. COPD Factor Analysis Axes (FAs): Previously published phenotype disease axes identified through factor analysis(19). 2. PCA Image Only Axes (PCA-Is): Disease axes constructed by applying Principal Component Analysis (PCA) to the CSRL features. The comparative analyses included the calculation of Pearson correlation coefficients between IEAs and other disease axes, association analyses to clinical measurements, and comparison of nested models for clinical outcomes utilizing IEAs, FAs, and PCA-Is in determining whether IEAs improved the performance of models already containing FAs and/or PCA-Is.

### Differential Expression and Usage Analyses

To identify the genes and pathways associated with IEAs, we performed differential gene expression analysis using limma and voom(20,21). Multiple comparisons were corrected with the Benjamini-Hochberg method to control the false discovery rate (FDR) at 10%(22). Gene Ontology (GO) pathway enrichment analysis was performed for pathways in the “Biological Process” category using the TopGO (v2.33.1) method (23). The threshold for statistical significance was an adjusted P-value < 0.001.

## Results

One thousand two hundred and twenty-three subjects in the COPDGene study with complete CT scan and blood RNA-seq data were analyzed, and the flow diagram for the selection of subjects for analysis is shown in Supplemental Figure E1. The analyzed subjects were 50% female, 82% non-Hispanic white, and 18% African American, and the average age of the subjects was 67 years. The GOLD spirometric stage distribution of subjects was 42.5% GOLD 0, 44.0% in GOLD 1-4, and 13.5% with preserved ratio impaired spirometry (PRISm). For model training and validation, subjects were split into training and test sets, with no statistically significant differences in demographic or key clinical characteristics between these groups (Table 1).

**Table 1.**
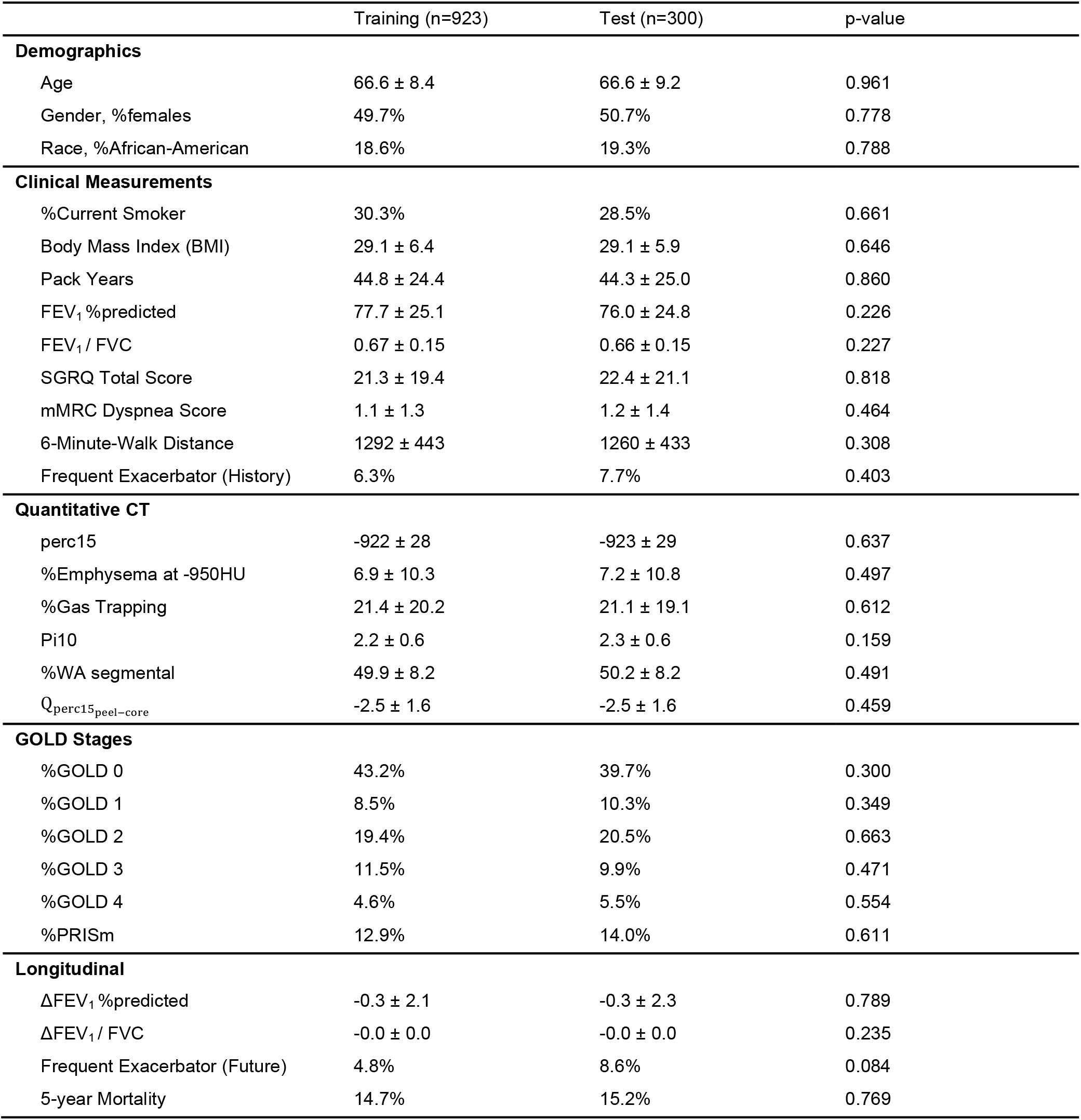
Subject characteristics in training and test data. Continuous variables are expressed as means and standard deviations. Categorical variables are expressed as percentages. P-values are obtained using the Kruskal-Wallis test for continuous variables and chi-square test for proportions, comparing the training and test data. *FEV*_1_ = Forced expiratory volume in 1 second; FVC = Forced vital capacity; perc15 = 15th Percentile Hounsfield unit in Inspiratory CT scan; %Gas Trapping: %LAA using −856 Hounsfield unit threshold on expiratory CT scan; Pi10 = the average wall thickness for a hypothetical airway of 10-mm lumen perimeter on CT; %WA segmental= the percentage of airway wall area for 3rd generation bronchi; GOLD: Global Initiative for Chronic Obstructive Lung Disease; PRISm=Preserved ratio impaired spirometry.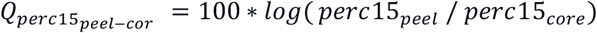, where the peel region is defined to be <5mm from the lung boundary and the core region is >20mm from the lung boundary. *ΔFEV*_1_ %*predicted* and *ΔFEV*_1_/*FVC* are computed by subtracting the visit 3 values from the visit 2 values of *FEV*_1_ % of predicted or *FEV*_1_/*FVC* and dividing them by the number of years between the two visits.

### IEA Model Training and Reproducibility Analysis

A schematic overview of the model training process is shown in Figure 1, and the data flow is summarized in Supplemental Figure E2. To maximize the stability of the IEAs and reduce the effects of sampling variability, we used nested cross-validation in the model training process to select the number of genes included in the model and the number of IEAs identified. For gene selection, we tested genes in the training data for the association to the top 128 principal components of the CSRL image features using an F-test, and a series of p-value thresholds for gene inclusion were explored, ranging from p = 1x10^−6^ to p = 1. The resulting IEAs were found to be stable across the entire range of p-value thresholds, and the threshold corresponding to p=0.01 was selected (Pearson’s r for IEAs across cross-validation folds ≥ 0.96, Supplemental Table E1). With this threshold, 4,685 genes were included in the final model. The number of IEAs identified by the model was determined by the amount of gene expression variance explained, which was the highest with two IEAs (Figure 2).

**Figure 1.**
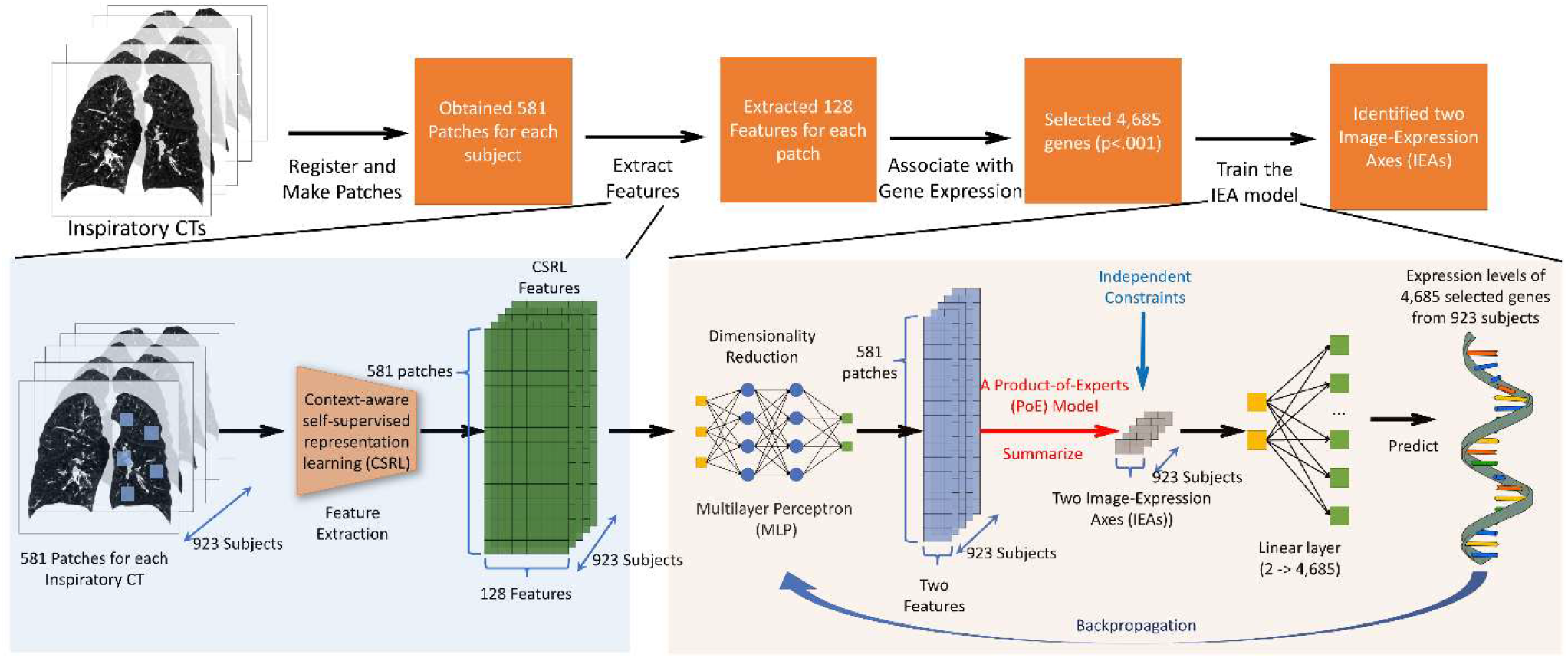
Overview of the machine learning workflow. CT images were processed as 32^3^ mm^3^ patches from which 128 features were constructed using the Context-Aware Self-supervised Representation learning algorithm (CSRL) (14). These features were the input for a multilayer perceptron (MLP) that processed a 581x128x923 tensor to output a 581x2x923 subject-level data representation. The subject-level latent representation (IEAs) is given by summarizing the patch-level features into a matrix of 2x923. We introduce a linear layer (2 -> 4,685) that estimates gene expression for each subject, taking the IEA as the input. We apply independent constraints to ensure IEAs are independent of each other. The overall objective function is given by minimizing the mean-squared error of the gene expression levels in prediction.

**Figure 2.**
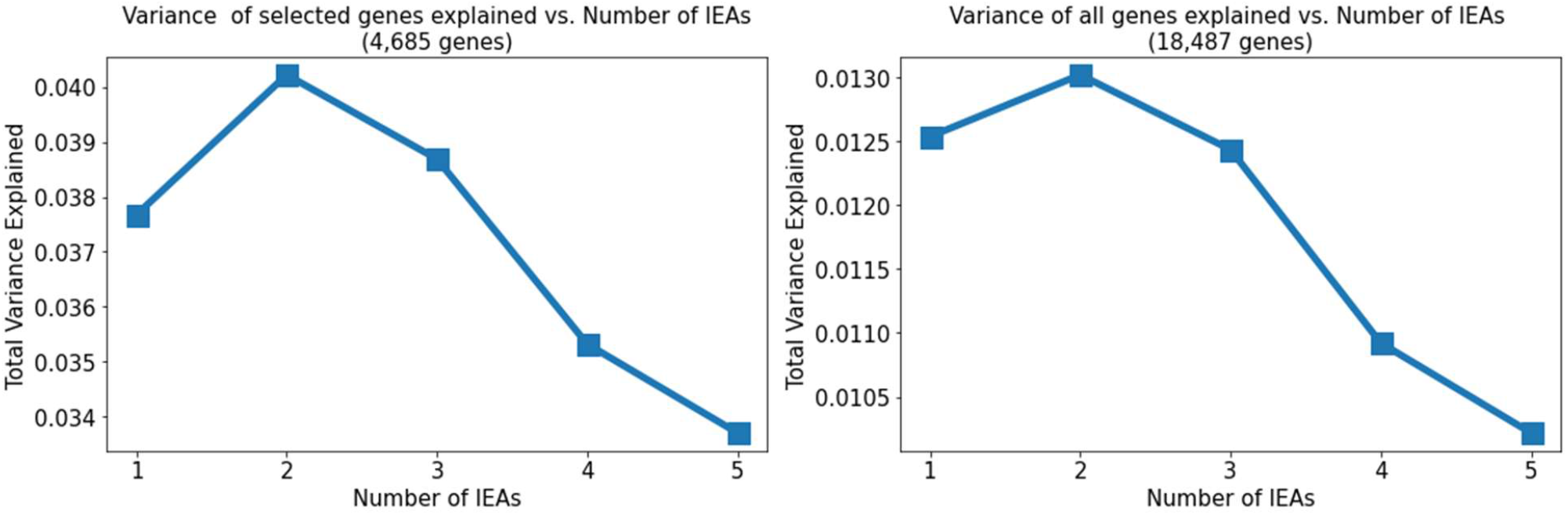
Total variance of the gene expression explained vs. number of IEAs. The figure on the left shows the plot of the 4,685 selected genes. The figure on the right shows the plot for all the genes. The figures show that when the number of IEAs is two, the total variance explained is maximized. We choose the number of IEAs to be two.

### IEA Association to Clinical and Radiographic Features and Prospective Outcomes

To provide a clinical interpretation of the IEAs, we calculated their correlation to a range of COPD-related clinical and imaging measurements (Table 2). We refer to the first IEA as the emphysema axis (IEA_emph_), because it demonstrates a pattern of clinical associations consistent with quantitative emphysema. Specifically, higher levels of IEA_emph_ were associated with lower lung function, emphysema, and lower BMI. The second IEA is consistent with airway disease and is referred to as IEA_airway_. Higher levels of IEA_airway_ were associated with higher BMI, thicker airways, and less emphysema. The IEAs were uncorrelated with each other, suggesting that they may capture different underlying disease processes.

**Table 2.** Pearson’s correlation between image-expression axes (IEAs) and COPD-related characteristics and health outcomes. The symbols “*”, “**”, and “***” represent p<.05, p<.01, and p<.001, respectively. FEV_1_ = Forced expiratory volume in 1 second; FVC = Forced vital capacity; perc15 = 15th Percentile Hounsfield unit in Inspiratory CT scan; %Gas Trapping: %LAA using −856 Hounsfield unit threshold on expiratory CT scan; Pi10 = the average wall thickness for a hypothetical airway of 10-mm lumen perimeter on CT; %WA segmental= the percentage of airway wall area for 3rd generation bronchi; 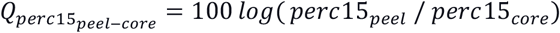, where the peel region is defined to be <5mm from the lung boundary and the core region is >20mm from the lung boundary; *ΔFEV*_1_ %predicted and *ΔFEV*_1_/*FVC* are computed by subtracting the visit 3 values from the visit 2 values of *FEV*_1_ %predicted or *FEV*_1_/*FVC* and dividing it by the number of years between the two visits.

|  | IEA<sub>emph</sub> | IEA<sub>airway</sub> |
| --- | --- | --- |
| **Clinical Measurements** |  |  |
| Body Mass Index (BMI) | -0.14* | 0.68*** |
| Pack Years | 0.28*** | 0.12* |
| FEV<sub>1</sub>%predicted | -0.45*** | 0.00 |
| FEV<sub>1</sub>/FVC | -0.65*** | 0.19*** |
| SGRQ Total Score | 0.40*** | 0.08 |
| mMRC Dyspnea Score | 0.40*** | 0.12* |
| 6-Minute-Walk Distance | -0.22*** | -0.30*** |
| **Quantitative CT** |  |  |
| perc15 | -0.76*** | 0.54*** |
| %Emphysema at -950HU | 0.72*** | -0.34*** |
| %Gas Trapping | 0.74*** | -0.32*** |
| Pi10 | 0.14* | 0.17** |
| %WA segmental | 0.09 | 0.34*** |
| $Q_{perc15_{peel-core}}$ | -0.36*** | -0.32*** |
| **Longitudinal** |  |  |
| $\Delta FEV_1$%predicted | -0.03 | 0.14 |
| $\Delta FEV_1$/FVC | -0.03 | 0.07 |
| **Image-Expression Axes (IEAs)** |  |  |
| IEA<sub>emph</sub> | 1.00*** | -0.03 |
| IEA<sub>airway</sub> | -0.03 | 1.00*** |

As shown in Table 2, both IEAs were negatively associated with emphysema peel/core distribution 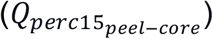, indicating that higher IEA values are associated with more emphysema in the core/central regions of the lung. To explore this association further, we performed sensitivity analyses, in which the lung was separated into concentric bands according to the distance to the lung boundary as described in the online supplement.

To determine whether the IEAs provided clinical information in addition to standard demographic variables, we tested the significance of adding IEAs to regression models for various COPD-related measures (Table 3, Table 4, and Supplemental Tables E2 and E3). After adjusting for standard demographic variables, both IEA_emph_ and IEA_airway_ were significantly associated with FEV_1_ %predicted, FEV_1_/FVC, SGRQ total score, mMRC dyspnea score, and 6-minute-walk distance. IEA_emph_ was also associated with Frequent Exacerbator (History), Frequent Exacerbator (Future), and overall mortality rate.

**Table 3.**
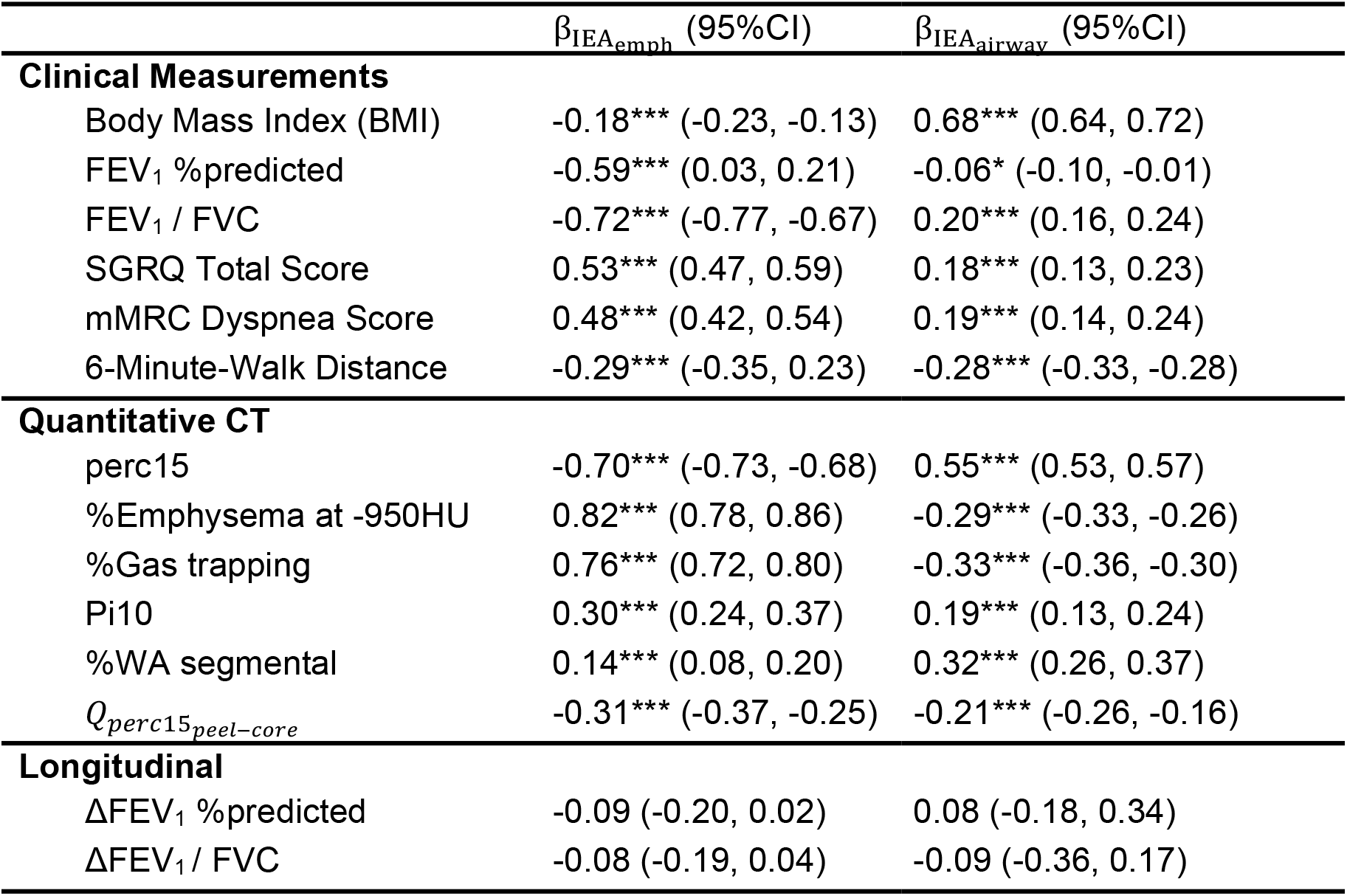
Multivariable associations of image-expression axes (IEAs) to continuous COPD-related characteristics and health outcomes. The table reports the β coefficients and corresponding 95% confidence intervals for IEA_emph_ and IEA_airway_ in linear models using the indicated COPD-related measurement or health outcomes as the response variable. All models were adjusted for age, gender, race, pack years, smoking status. The symbols “*”, “**”, and “***” represent p<.05, p<.01 and p<.001, respectively. *FEV*_1_ = Forced expiratory volume in 1 second; FVC = Forced vital capacity; perc15 = 15th Percentile Hounsfield unit in Inspiratory CT scan; %Gas Trapping: %LAA using −856 Hounsfield unit threshold on expiratory CT scan; Pi10 = the average wall thickness for a hypothetical airway of 10-mm lumen perimeter on CT; %WA segmental= the percentage of airway wall area for 3rd generation bronchi; 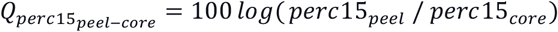, where the peel region is defined to be <5mm from the lung boundary and the core region is >20mm from the lung boundary. *ΔFEV*_1_ %*predicted* and *ΔFEV*_1_/*FVC* are computed by subtracting the visit 3 value from the visit 2 value of *FEV*_1_ % *of predicted* or *FEV*_1_/*FVC* and dividing it by the number of years between the two visits.

**Table 4.** Multivariable associations of image-expression axes (IEAs) to frequent exacerbations and mortality. The table reports the β coefficients of the IEA_emph_ and IEA_airway_ and corresponding 95% confidence intervals from logistic regression models for frequent exacerbator status and a Cox proportional hazards model for mortality. All models adjusted for age, gender, race, pack years, smoking status as the covariates. The symbols “*”, “**”, and “***” represent p<.05, p<.01, and p<.001, respectively. Frequent exacerbator (history) indicated whether the subject had at least two self-reported exacerbations during the 12 months before the second visit. Frequent exacerbator (future) indicated whether the subject had at least two self-reported exacerbations during the past 12 months before the third visit.

| **Logistic Regression Model** | $\beta_{IEA_{emph}}$<br>(95%CI) | Odds Ratio<br>(95%CI) | $\beta_{IEA_{airway}}$<br>(95%CI) | Odds Ratio<br>(95%CI) |
| --- | --- | --- | --- | --- |
| Frequent Exacerbator<br>(History) | 0.85***<br>(0.58, 1.12) | 2.34***<br>(1.79, 3.07) | 0.12<br>(-0.10, 0.35) | 1.13<br>(0.90, 1.41) |
| Frequent Exacerbator<br>(Future) | 0.92***<br>(0.47, 1.37) | 2.51***<br>(1.60, 3.94) | -0.09<br>(-0.48, 0.30) | 0.91<br>(0.62, 1.34) |

| **Cox Proportional Hazard Model** | $\beta_{IEA_{emph}}$<br>(95%CI) | Hazard Ratio<br>(95%CI) | $\beta_{IEA_{airway}}$<br>(95%CI) | Hazard Ratio<br>(95%CI) |
| --- | --- | --- | --- | --- |
| Mortality | 0.66***<br>(0.45, 0.87) | 1.93***<br>(1.57, 2.38) | 0.17<br>(-0.00, 0.34) | 1.19<br>(1.00, 1.41) |

To provide independent replication of our IEA associations, the IEA model was applied to 1,527 subjects from another subset of the COPDGene dataset that had not been used for model training. All the significant associations to clinical and longitudinal measures remained significant with very similar effect estimates indicating a high level of reproducibility for IEAs (Supplemental Tables E4, E5 and E6).

### COPD Subgroups Defined by IEAs and Comparison to Existing COPD Subtypes

To further understand the clinical characteristics of COPD subgroups defined by IEAs, we divided the IEA space into four quadrants (Figure 3) and computed the average characteristics of each subgroup (Supplemental Tables E7). As expected, subjects with low IEA_emph_/low IEA_airway_ values had the least obstruction (mean FEV1 89.2% predicted), the highest percentage of GOLD spirometric grade 0 subjects, low emphysema, and the thinnest airways. Subjects with high IEA_emph_/low IEA_airway_ values had characteristics consistent with emphysema-predominant COPD, namely high emphysema and low BMI, with about 70% of GOLD grade 4 subjects present in this group. Subjects with low IEA_emph_/high IEA_airway_ values had an airway-predominant profile with thick airway walls, elevated BMI, and the greatest proportion of PRISm subjects. Subjects with high IEA_emph_/high IEA_airway_ had the highest SGRQ total score, highest mMRC dyspnea scores, and shortest 6-minute walk distance. In terms of COPD progression, the groups differed significantly in mortality risk (p<0.001) and frequent exacerbation status (2 or more exacerbations in one year) but did not change in FEV1. The group with the highest mortality was high IEA_emph_/high IEA_airway_ followed by high IEA_emph_/low IEA_airway_. The latter group also had the highest percentage of subjects with frequent exacerbations, both for retrospective (p<0.001, chi-square test of all groups) and prospective exacerbations (p=0.048).

**Figure 3.**
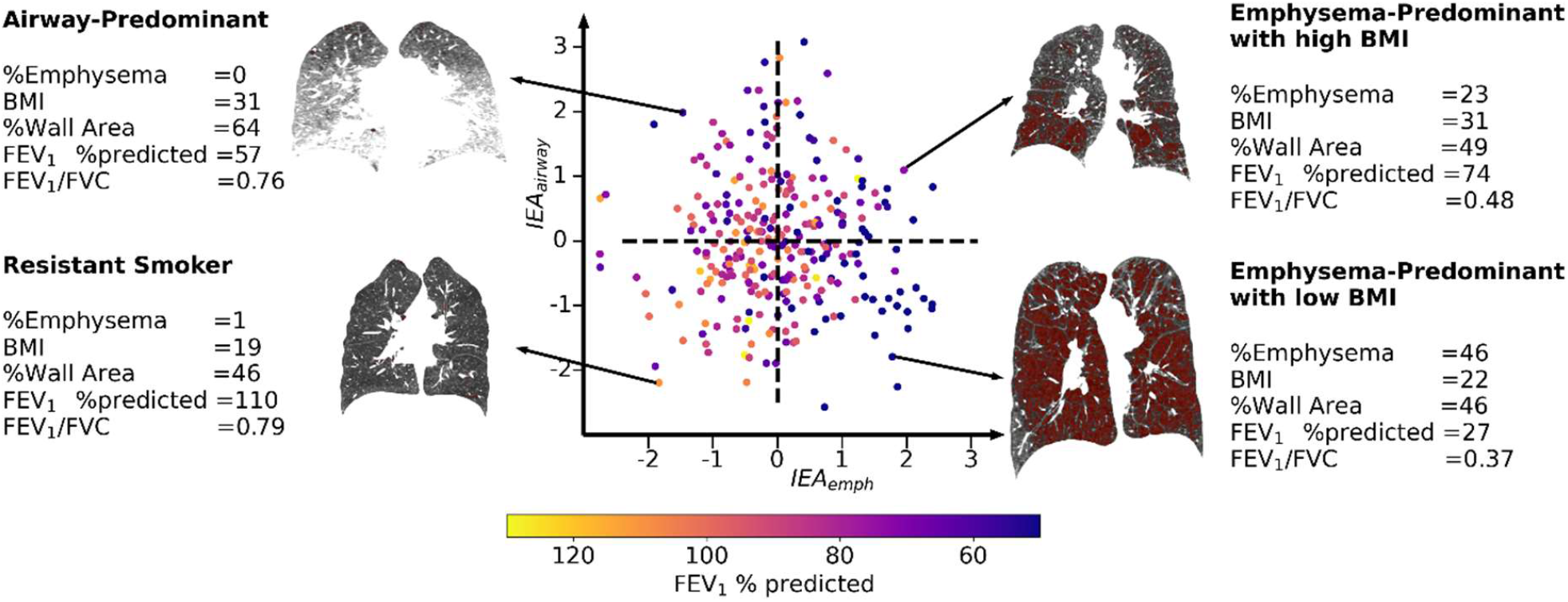
Visualization of subjects projected along each identified image-expression axis (IEA) dimension. IEA_emph_ is the emphysema axis, where higher values indicate more severe emphysema. IEA_airway_ is the airway disease axis, where a higher value represents higher BMI and thicker airways. The space defined by these IEAs was used to stratify the cohort into four subgroups based on dividing the IEA space into four quadrants. Lung CT scans and clinical characteristics are shown for one subject in each quadrant, where the red mask represents the emphysema regions (< -950 HU). The characteristics of the four quadrants are summarized in Supplemental Figure E5.

To place IEA_emph_ and IEA_airway_ in the context with previously reported subtypes and disease axes in COPDGene, we compared these axes directly with the previously reported k-means subtypes (24) and factor-analysis derived disease axes (FAs) (19). In Figure 4, we observe that the highest values of IEA_emph_ are found in the severe emphysema k-means subtype, and the highest values of IEA_airway_ are found in the airway-predominant k-means subtype, confirming our clinical interpretation of these disease axes. Since the FAs also showed patterns consistent with emphysema (FA_emph_) and airway-predominant disease (FA_airway_), we compared the IEAs to the FAs and observed that IEA_emph_ and FA_emph_ showed a reasonably strong correlation (Pearson’s r = 0.58), but the IEA_airway_ and FA_airway_ axes showed only modest correlation (Pearson’s r = 0.28, see Supplemental Table E8). Examination of the pattern of clinical associations for IEA_airway_ and FA_airway_ revealed that while airway axes were positively correlated to airway wall thickness, IEA_airway_ is negatively correlated to emphysema whereas FA_airway_ is positively correlated (Supplemental Table E9). To determine whether the IEAs provided additional information about COPD phenotypes (FEV1 % of predicted, FEV1/FVC, SGRQ, mMRC, retrospective frequent exacerbations, and 6-minute walk distance) and COPD progression (mortality and prospective frequent exacerbations) above and beyond FAs, we constructed baseline models for each COPD phenotype and progression measurements with FAs included and then compared them to models including both FAs and IEAs. In most cases the models with IEAs included outperformed the baseline models (p<0.001 for all COPD phenotypes and mortality, Supplemental Tables E10 and E11).

**Figure 4.**
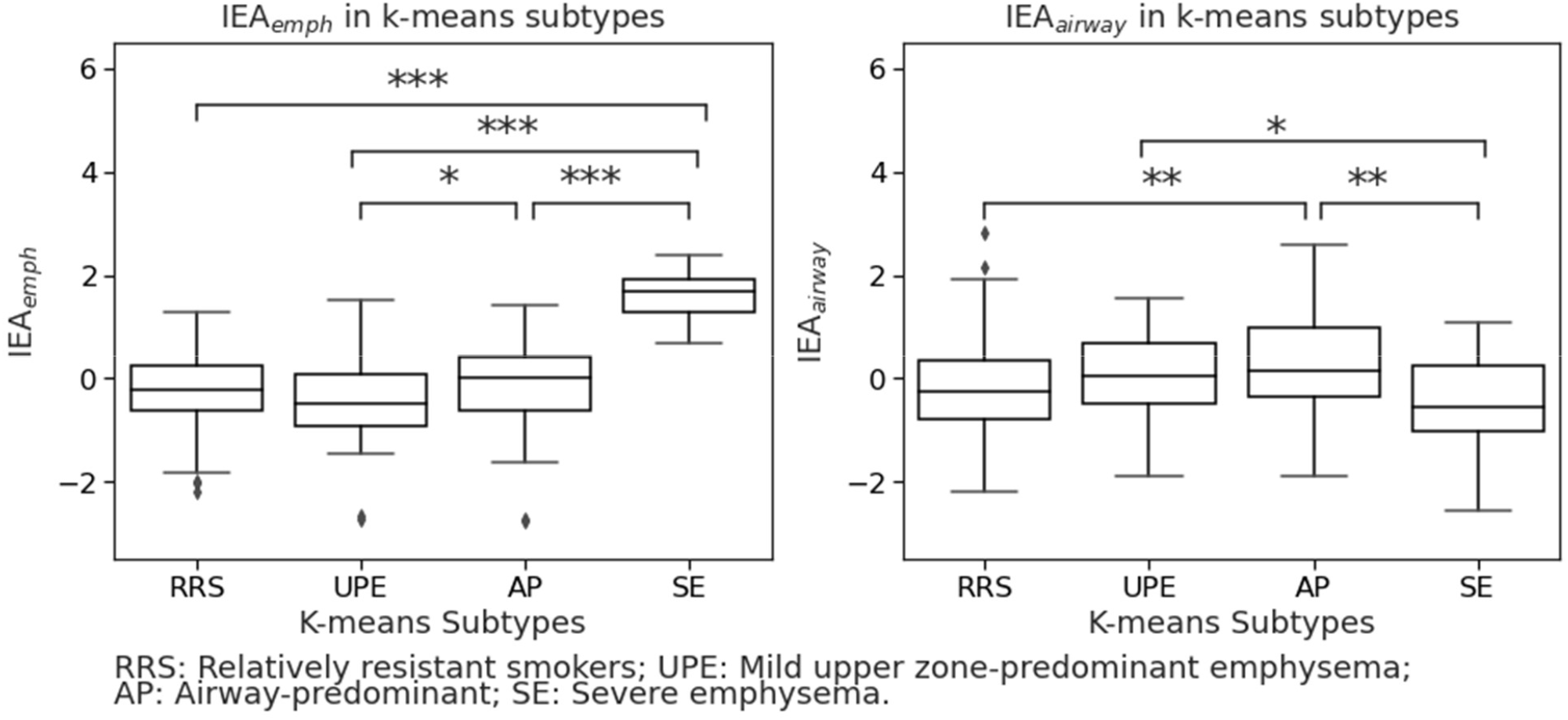
Distribution of IEA_emph_ and IEA_airway_ values grouped by previously published COPD K-means clustering subtypes(24). P-values are obtained using the Kruskal-Wallis test. The symbols “*”, “**”, and “***” represent p<.05, p<.01, and p<.001, respectively, for a t-test comparing to the Relatively Resistant Smokers Group. No symbol indicates non-significant results.

### Comparison to PCs based on Images Alone

After observing that IEAs contain additional clinically relevant information relative to standard features extracted from CT images, we sought to determine whether the additional information came only from applying dimension reduction to the CT images (CSRL features), or whether there was added value from our algorithm that combined the CT features with gene expression. To make this comparison, we constructed disease axes from images only by using PCA to extract the top-2 PCs of the CSRL features, denoted as PCA-Is. We then compared the predictive performance of linear models that utilize both IEAs and PCA-Is with the nested version that involves the PCA-Is only, and we observed that the models including IEAs were superior to models with PCA-Is only for all of the six studied COPD phenotypes as well as prospective exacerbations and mortality (p<0.001, Supplemental Tables E12 and E13). These results suggest that by incorporating gene expression data during training, IEAs extract more clinically important information than similar methods that utilize imaging features only.

### IEAs are associated with inflammatory pathways

To understand the biological aspect of the IEAs, we first confirmed that IEAs explained a greater proportion of gene expression variance that PCA-Is, as demonstrated in Figure 5, which shows that IEAs explain a greater proportion of variation on a per-gene basis than PCA-Is (557 genes with R^2^ >10% for IEAs versus 68 genes with R^2^ >10% for PCA-Is).

**Figure 5.**
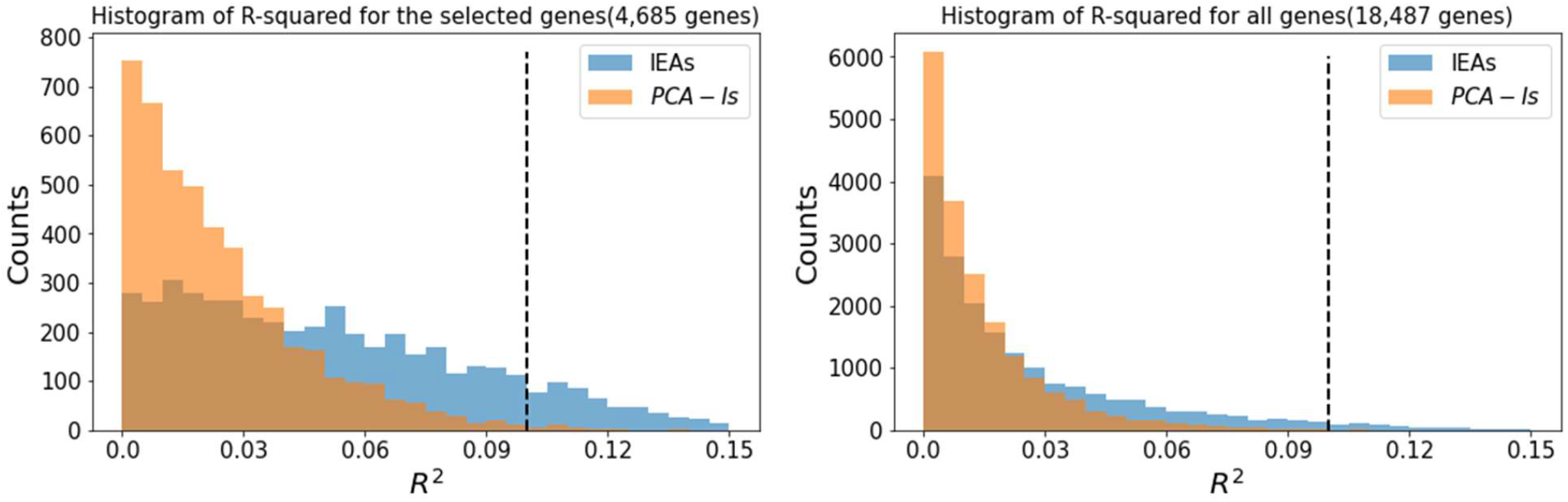
Histograms for the variances of genes explained (R^2^) by the image-expression axes (IEAs) and PCA Image Only Axes (PCA-Is). The figure on the left shows the histogram for the 4,685 selected genes. Within these genes, there are 557 genes with R^2^ >10% for IEAs and 68 genes with R^2^ >10% for PCA-Is. The figure on the right shows the histogram for all the genes (18,487 genes in total). There are 622 genes with R^2^ >10% for IEAs and 69 genes with R^2^ >10% for PCA-Is.

To identify specific biological processes associated with each IEA, we performed differential expression and pathway enrichment analysis. We identified 6,494 and 3,815 genes associated at an FDR of 10% with IEA_emph_ and IEA_airway_, respectively (Supplemental Tables E14 and E15). Gene Ontology pathway enrichment identified 29 and 13 enriched pathways (p-value < 0.001) for IEA_emph_ and IEA_airway_, respectively (Supplemental Tables E16 and E17). The most significantly associated pathway for IEA_emph_ was neutrophil degranulation, whereas IEA_airway_ had the strongest enrichment for RNA processing. The most significant pathway results are shown in Table 5.

**Table 5.** Top-10 Significant Gene Ontology enrichment terms for IEAs. Gene ontology (GO) pathway enrichment analysis was performed using the GO “Biological Process” gene sets with p-values calculated with the Fisher exact test statistic using the weight01 algorithm in topGO(*23*) (v2.33.1) that accounts for dependency in GO topology.

**Top-10 Significant Gene Ontology enrichment terms for IEA<sub>emph</sub>**
| GO terms | Description | Annotated | Significant | adjusted p-values |
| --- | --- | --- | --- | --- |
| GO:0043312 | neutrophil degranulation | 461 | 239 | 2.60e-18 |
| GO:0019083 | viral transcription | 174 | 99 | 4.20e-17 |
| GO:0000184 | nuclear-transcribed mRNA catabolic process, nonsense-mediated | 119 | 79 | 2.00e-14 |
| GO:0006614 | SRP-dependent cotranslational protein targeting to membrane | 99 | 65 | 2.20e-13 |
| GO:0006413 | translational initiation | 185 | 93 | 2.30e-13 |
| GO:0002181 | cytoplasmic translation | 98 | 51 | 2.20e-07 |
| GO:0006364 | rRNA processing | 224 | 124 | 6.40e-07 |
| GO:0006954 | inflammatory response | 529 | 250 | 4.40e-06 |
| GO:0006955 | immune response | 1758 | 750 | 2.00e-05 |
| GO:0045730 | respiratory burst | 33 | 20 | 3.30e-05 |

| GO terms | Description | Annotated | Significant | adjusted p-values |
| --- | --- | --- | --- | --- |
| GO:0006396 | RNA processing | 981 | 204 | 1.80e-08 |
| GO:0098869 | cellular oxidant detoxification | 77 | 33 | 1.00e-06 |
| GO:0042744 | hydrogen peroxide catabolic process | 24 | 14 | 1.70e-05 |
| GO:0006879 | cellular iron ion homeostasis | 56 | 25 | 7.30e-05 |
| GO:0001895 | retina homeostasis | 43 | 16 | 9.10e-05 |
| GO:0030449 | regulation of complement activation | 79 | 30 | 9.20e-05 |
| GO:0019886 | antigen processing and presentation of exogenous peptide antigen via MHC class II | 90 | 32 | 1.00e-04 |
| GO:0006413 | translational initiation | 185 | 53 | 1.00e-04 |
| GO:0006614 | SRP-dependent cotranslational protein targeting to membrane | 99 | 34 | 2.40e-04 |
| GO:0050900 | leukocyte migration | 377 | 89 | 5.20e-04 |

## Discussion

In this paper, we used deep learning to identify novel connections between lung imaging features and blood gene expression. The deep learning model provided novel disease axes, i.e. IEAs, that captured elements of shared variability between CT scans and blood RNA-seq, and we demonstrated 1) that these IEAs are associated with important COPD-related physiologic and functional measures, 2) that these associations contained information that is independent from pre-existing, standard clinical and imaging variables, 3) that the IEA_emph_ axis is significantly associated with prospective mortality in multivariable models, and 4) that IEAs capture distinct patterns of connection between lung structural changes and systemic inflammation.

Many of our main results are consistent with our current understanding of COPD. IEAs capture the two cardinal pathologies of COPD, emphysema, and airway disease; but clearer links between these aspects of lung structure and systemic inflammation emerge from the joint analysis of CT images and blood RNA-seq. First, neutrophilic inflammation was strongly associated with emphysema but not the airway axis. This agrees with the prominent role of neutrophils in alpha-1 antitrypsin associated (AAT) emphysema(25), and it provides further support for the role of neutrophils in the emphysema of “typical” COPD. There is evidence as well for a role for specific adaptive immune processes that show significant enrichment for both IEA_emph_ and IEA_airway_, though the inflammatory signals that we observed in blood differ from the B-cell predominated signatures that have been observed in some lung transcriptomic studies of emphysema(26). The biological pathway enrichments we observed are consistent with previous reports of the association of emphysema to biomarkers related to systemic inflammation, oxidative stress, and elevated plasma fibrinogen levels(27). IEA_airway_ is strongly correlated to BMI, which coincides with a previous hypothesis that obesity-related adipose tissue hypoxia and systemic hypoxia due to reduced pulmonary function contribute to the systemic inflammation of COPD(28). Future studies, including single cell transcriptomic data, could better identify the association of emphysema and airway disease with specific types of innate and adaptive inflammatory cells.

While our IEAs seem most descriptive of emphysema and airway disease, they are not completely correlated to standard CT measurements of emphysema and airway disease, and they differ notably from machine-learning disease axes based on imaging alone (PCA-Is) or based on imaging and spirometry (FAs) (19). While none of these representations of emphysema and airway disease is demonstrably superior to the others, the IEA axes have a clear interpretation due to the integrative nature of the deep learning algorithm, whose goal was to find shared variability between CT images and transcriptomic patterns in the blood. The clinical relevance of these axes was demonstrated through regression models showing that IEAs were significantly associated with a wide range of COPD-related measures, including mortality. In the future, these algorithms can be extended to incorporate additional sources of molecular or imaging data.

While the IEAs primarily captured patterns of emphysema and airway-predominant COPD, they were also significantly correlated with core versus the periphery (“peel”) emphysema distribution. Previous work has demonstrated numerous clinically relevant associations to aspects of emphysema distribution, most notably for core/peel and apical/basal emphysema distribution(29-33), and a machine learning analysis of images alone also identified peel-core emphysema distribution as an important dimension of COPD-related variability(34). Our analysis suggests that systemic inflammation is most strongly associated with core/peel rather than apical/basal emphysema distribution, and that the amount of emphysema in the core region is associated with the more severe disease along both the IEA_emph_ and IEA_airway_ axes. Since quantification of the lung peel can be influenced by technical factors related to lung segmentation, we conducted sensitivity analyses that confirmed a consistent association for the IEA_emph_ axis, whereas the IEA_airway_ association was clearly present only when the analysis included the outermost lung regions. Accordingly, we have high confidence in the IEA_emph_ association to core/peel distribution, but it is not clear whether the IEA_airway_ axis association reflects a true biological relationship or technical artifacts.

By collecting CT scans, blood transcriptomics, and detailed phenotype data on thousands of current and former smokers enriched for COPD, the COPDGene Study provides a novel opportunity for the application of machine learning to better understand the connections between the lung structure in COPD and molecular mechanisms of systemic inflammation. Like all machine learning models, the construction of our model required many explicit and implicit design choices. In our model, we extracted patch-level representations via self-supervised learning. Such methods are capable of extracting generalized and semantically meaningful features(35). The linear independence assumption in our model was intended to identify IEAs that captured distinct underlying disease processes and increase the reproducibility of our model. Unlike previous studies that explore the relationship between COPD imaging and omics by associating previously discovered image patterns to omics data(24,36,37), our method potentially identifies new image patterns that have not been previously explored.

The main strengths of this study are: 1) The joint analysis of full DICOM data from CT images and gene expression data via deep learning is novel and provides new biological and clinical insight into COPD. 2) The sample size is large, allowing for more power to identify novel discoveries. 3) We used a number of techniques to improve the reproducibility of our disease axes, including cross-validation, sensitivity analysis, and the use of constraints in our modeling procedure. The main limitations are: 1) Our study is limited to blood RNA biomarkers, which capture systemic inflammation but no other important aspects of the COPD inflammatory response, such as lung gene expression and protein biomarkers. 2) Our analysis was limited to the COPDGene study. In the future, such analyses could be conducted in other ongoing studies collecting CT scan and omics data in populations enriched for COPD.

In summary, deep learning applied to CT images, and transcriptomic biomarkers in COPD identified two main inflammatory processes related to CT image features that can be broadly defined as emphysema and airway disease. The emphysema-related process was most enriched for pathways related to neutrophilic inflammation. The airway axis differed from previously reported disease axes learned from phenotypic data alone and it was negatively correlated with emphysema. Finally, there was also a strong relationship between the core-peel distribution of emphysema and systemic inflammation. In the future, these integrative machine learning methods can be refined for more fine-grained interpretability and extended to include other sources of biological information.

## Data Availability

All data produced are available online at dbGaP

https://www.ncbi.nlm.nih.gov/projects/gap/cgi-bin/study.cgi?study_id=phs000179.v6.p2

## Supplemental Methods

### Subject Enrollment and Data Collection

COPDGene enrolled 10,198 subjects with a minimum 10 pack-years lifetime smoking history at 21 centers across the United States (NCT00608764, www.copdgene.org)(1). Five-year follow-up data are available for 6,717 subjects, and 10-year follow-up visits are currently being completed. Subjects underwent spirometry, questionnaire assessments, standardized inspiratory and expiratory chest CT imaging, and genome-wide genotyping. Starting at the second visit (year 5), PAXgene blood RNA tubes were collected. Each center obtained institutional review board approval, and all participants provided written informed consent.

We used the following cross-sectional covariates from the COPDGene second study visit in our analysis – age, race, gender, postbronchodilator spirometric measures, Body Mass Index (BMI), St. George’s Respiratory Questionnaire (SGRQ) total score, modified Medical Research Council (mMRC) Dyspnea Scale, and the 6-minute walk distance The variable “frequent exacerbator (history)” indicates whether the subject had >=2 self-reported exacerbations during the 12 months before the second visit. Exacerbations were defined as worsening respiratory symptoms treated with systemic steroids and/or antibiotics.

The following quantitative CT measurements were analyzed: Perc15 (15th percentile value of Hounsfield unit (HU) in the lung region), % emphysema (the percentage of low attenuation areas less than -950 HU in the inspiratory CT scan), % air trapping (the percentage of low attenuation areas less than -856 HU in the expiratory CT scan), Pi10 (The square root of the wall area of a theoretical airway with an internal lumen perimeter of 10 mm) and %WA segmental (the percentage of airway wall area for 3rd generation bronchi). The variable 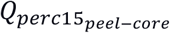 is defined as 100 times the logarithm of the ratio of perc15 value in the peel to the core region of the lung. We define the peel region as the lung region that is not more than 5mm from the boundaries of the lung and the core region to be more than 20mm from the lung boundary. A larger value of 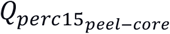 represents that more emphysema is in the peel area of the lung.

Using data gathered from a subset of subjects who had already completed the third (10-year) COPDGene Study visit, we included variables on prospective change in lung function and exacerbations in our analysis. ΔFEV_1_ %predicted and ΔFEV_1_/FVC are computed by subtracting the visit 3 value from the visit 2 value of FEV_1_ or FEV_1_/FVC and dividing it by the number of years between the two visits. We used “frequent exacerbator (future)” to represent whether the subject had at least two self-reported exacerbations during the 12 months prior to the third study visit.

### Extracting CSRL Features from CT Scans

We resampled inspiratory CT scans to isotropic 1mm^3^ resolution. The Hounsfield Units (HU) were mapped to the intensity window of [−1024, 240]. We selected the CT scan from a healthy subject as the anatomical atlas. Next, we divided the atlas image into non-overlapping patches with the size 32^3^ mm^3^ using the sliding window approach, which resulted in 581 patches. The center locations of these patches were used as anatomical landmarks. Then we performed affine registration using ANTs (2) to map these atlas landmark coordinates to every CT scan, which maintained the anatomical correspondences between images. Finally, we divided each CT scan into 581 patches centering at the mapped anatomical landmarks, with a size of 32^3^ mm^3^. We extracted the features for each patch using the context-aware self-supervised representation learning (CSRL) model (3). The CSRL model was pre-trained using the inspiratory CT scans collected during the first COPDGene study visit. This model embedded each patch in 128-dimensional space, resulting in 128 CSRL features, assuming the embedding of similar samples are closer and diverse samples are far from each other.

### RNA-seq Data Generation and pre-processing

Total blood RNA was collected at Visit 2 in PAXgene TM Blood RNA tubes from the Qiagen PreAnalytiX PAXgene Blood miRNA Kit (Qiagen, Valencia, CA). Paired-end reads were generated from Illumina sequencers. Gene transfer format (GTF) annotation was downloaded from the Biomart Ensembl database (Ensembl Genes release 94, GRCh38.p12 assembly).

Gene counts were derived from Salmon isoform estimates summarized at the gene level using the *tximport* package. The gene and isoform count data used for this analysis are available in the Gene Expression Omnibus(4) (accession number GSE158699).

Genomic features with very low expression (average counts per million (CPM) < 0.2 or number of subjects with CPM < 0.5 was < 50) or extremely highly expressed genes (number of subjects with CPM > 50,000 was less than 50) were filtered out prior to applying trimmed mean of M values normalization from edgeR (v3.24.3), which accounts for differences in sequencing depth(5). Counts were also transformed to log2 CPM values and quantile-normalized to further remove systematic noise from the data.

### Learning Image-Expression Axes (IEAs)

To select the most relevant genes to use in our machine learning modeling, we first concatenated 128 CSRL features in 581 patches for each subject into a 581× 128 vector and conducted Principal Component Analysis (PCA) to reduce the dimensionality to 128. Then we trained a linear regression model with these 128 Principal Components (PCs) to predict each gene expression level and computed the p-value for each regression model using an F-test. We corrected for multiple comparisons using the *Benjamini–Hochberg* procedure(6), and for the final model, we included genes with an adjusted p-value <0.01 based on sensitivity analyses described below. Gene expression data were centered and scaled prior to analysis (mean 0 and unit variance).

We implemented a machine learning model in PyTorch (v1.10.0) to extract Image-Expression Axes (IEAs) for each subject, utilizing the CSRL features of 581 patches for each subject. We first fed CSRL features to a multilayer perceptron (MLP) to obtain a low-dimensional representation for each patch. Then we obtained the subject-level disease axes, which we call Image-Expression Axes (IEAs), by summarizing the 581 sets of low-dimensional patch-level representations using a Product of Expert (PoE) model(7). The PoE model assumes the probability density functions (PDF) of the subject-level disease axes are given by the product of PDFs of the patch-level representations. We apply independent constraints for the IEA with the Hilbert-Schmidt independence criterion (HSIC) (8) to ensure statistical independence of each identified axis with the goal of each axis capturing an independent disease process. We added another linear layer that took the IEAs as the input and outputted the estimated expression levels of all selected genes simultaneously. We define the objective function of the model to minimize the mean-squared errors between the estimated and the observed expression levels of the selected genes. The parameters of the model were optimized via Adam(9). We include the details of the deep learning method in the next section.

### The Deep Learning Model

The context-aware self-supervised representation learning (CSRL) extracted 128 features for each of the 581 patches. We denote these CSRL features using a vector **x**_*n,p*_ ∈ **R**^128^, where *n* ∈ {1, …, *M*}, and *p* ∈ {1, …, 581} represent the indices for subjects and patches, respectively, and *M* is the number of subjects in the training set. We first learn an *L*-dimensional patch-level representation for each patch **r**_*n,p*_ ∈ **R**^L^. We assume that each element of **r**_*n,p*_ follows a Gaussian distribution. We assume that the prior distribution for each element of **r**_*n,p*_ is given by the standard Gaussian distribution with zero mean and unit variance such that p(r_*n,p,l*_) = *N* (0, 1), and its posterior can be approximated by 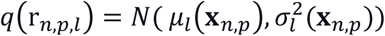, where *l* ∈ {1, …, *L*} is the index for the representation. The mean and variance of the Gaussian distribution *q*(r_*n,p,l*_), denoted as *μ*_l_(*·*) and 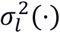 are the output of Multi-Layer Perceptron (MLP) with 3 hidden layers. We let the three hidden layers of the MLP contain 96, 64 and 48 nodes, respectively, with Rectified Linear Unit (ReLU) activation functions. We use ***θ*** to represent all the parameters for this MLP.

We summarize the patch-level representations into a subject-level IEA **z**_*n*_ ∈ **R**^L^ with a product-of-expert model(7), assuming that the distribution of each dimension in **z**_*n*_ is given by 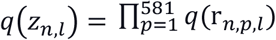. Since *z*_*n,l*_ is a produce of Gaussian distributions, *z*_*n,l*_ also follows a Gaussian distribution, such that 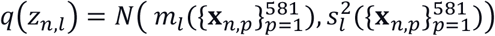, where

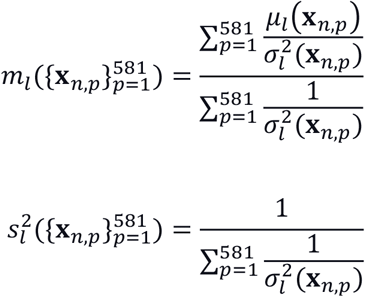

Note that both *m*_*l*_(*·*) and 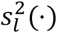 are functions of 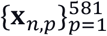, and parameterized by the MLP parameters ***θ***.

We assume that each dimension in **z**_*n*_ is independent with each other, such that each *z*_*n,l*_ represents an independent disease process of COPD. Therefore, we introduce a pairwise Hilbert-Schmidt independence criterion (HSIC) penalty in the objective function, such that *z*_*n,i*_ and *z*_*n,j*_ are statistically independent for all *i, j* ∈ {1, …, *L*}, denoted by 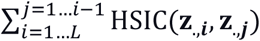,where HSIC is estimated as described in Section 5.2 in (8).

We denote the gene expression levels for each subject is given as **y**_*n*_ ∈ **R**^D^, where D represents the number of genes. We estimate **y**_*n*_ with a linear layer using **z**_*n*_ as the input, such that the estimator is given as **ŷ**_*n*_ *=* **Wz**_*n*_, where **W** ∈ **R**^D×L^ represent the parameters in the linear model. We define the objective function of the model to minimize the mean-squared errors between the estimated and the observed expression levels of the selected genes.

We optimize the evidence lower bound (ELBO)(10) as the objective function, which can be given as

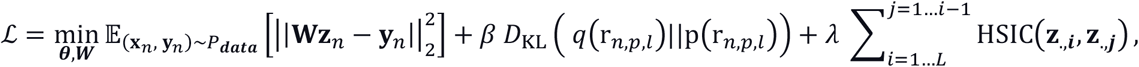

where 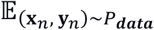 represent the expected values are estimated with the observed data, and *D*_*KL*_(*·* || *·*) represents the Kullback–Leibler. The hyper-parameters *β* and *λ* control the trade-off between the contributions of different terms in the objective function. In the primary experiments we let *β* = 0.1 and *λ* = 1.

We minimized the objective function utilizing Adaptive Moment Estimation (Adam) (9). In the optimization, we utilized the reparameterization trick, i.e., we let 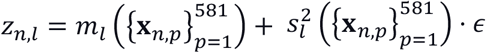 for each *n* ∈ {1, … *N*} and *l* ∈ {1, … *L*}, where *∈* is a random sample from *𝒩*(0, 1).

### Model training and Validation

We analyzed the inspiratory CT scans and the RNA-seq data of 1,223 subjects. We randomly split the data into training and testing sets containing 923 and 300 subjects, respectively. Model training was performed in the training set of 923 subjects using five-fold cross-validation, giving us five models. We aligned the IEAs by pairing the IEA from different models, such that the Pearson correlation coefficient is maximized. We standardize the aligned IEAs of each model, such that they have zero mean and unit variance. The final IEAs were given by taking the average value of the aligned IEAs given by five models.

We first validated our model by examining whether the learned IEAs were reproducible if different subsets of samples were used for training during the 5-fold cross-validation. We quantified the reproducibility via computing the mean and standard deviations of the Pearson correlation coefficients between the aligned axes. Then, we examined whether the IEAs were reproducible if different subsets of genes were used for training. To do so, we varied the thresholds for the gene selection process such that different subsets of genes were selected, conducted a 5-fold cross-validation, and obtained the final IEA by taking the average value of the standardized aligned IEAs of 5 folds. We compared these final IEAs trained with different subsets of genes by computing the Pearson correlation coefficients.

### Association of IEAs with Clinical Measurements

To understand the association between the IEAs and the clinical measurements, we conducted a univariable analysis by computing Pearson correlation coefficients between each of the IEAs and each of the clinical measurements. We conducted multivariable analysis via training an Ordinary Least Squares (OLS) model for continuous measurements and training a logistic regression model for categorical measurements. We conducted a survival analysis with the Cox proportional hazards model(11). In these multivariable analyses, we adjusted for the following covariates: age, race, gender, pack-years of smoking, current smoking status for all models. To determine whether the IEAs improved the prediction of each outcome, we compare the models with both IEAs as predictors to a reduced model without the IEAs using likelihood-ratio tests.

In this study, we utilize 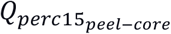 to measure the peel-core distribution of emphysema. To better understand how the peel-core distribution of emphysema is related to the IEAs, we conducted sensitivity analyses where we separated the lung into bands according to the distance to the lung boundary. We compute logarithm the ratio of perc15 value in different bands to the core region of the lung and observe how it is correlated to the IEAs.

To compare IEAs to previously identified COPD subtypes using k-means (12), we investigated whether the IEA values are different across the identified subtypes with the student’s t-test.

### Comparison of IEAs to Other COPD Subtypes and Disease Axes

We compared the IEAs to the following disease axes obtained with machine learning approaches: 1. COPD Factor Analysis Axes (FAs): Previously published phenotype disease axes identified through factor analysis of quantitative CT measurements and pulmonary function variables(13). 2. PCA Image Only Axes (PCA-I): Disease axes are constructed from imaging data only by applying Principal Component Analysis (PCA) to the same CSRL features used in the IEA analysis, but without using the gene expression data. Since the FAs were available only at the COPDGene first study visit, we generated IEAs from the CT scan data at visit 1 in order to directly compare FAs and IEAs. The comparative analyses were: 1. Calculating the Pearson correlation coefficients between IEAs and other disease axes; 2. Association analyses of FAs and PCA-I disease axes to the same cross-sectional and longitudinal clinical measurements described above; 3. likelihood-ratio tests to compare models utilizing both IEAs and FAs or PCA-I axes to baseline models including either FAs or PCA-I without IEAs.

To understand whether the IEAs capture gene-relevant information, we quantified the proportion of variation of gene expression explained by the IEAs in the COPDGene visit 2 data. We compared this to the proportion of expression variance explained by the PCs of CSRL features (PCA-I) to understand whether the IEAs capture more gene-relevant information.

### Differential Expression and Usage Analyses

To identify the genes and pathways associated with each of the IEAs identified in this study, we performed differential gene expression using the limma R package (v3.38.3)(14,15). We adjusted for the following covariates: age, race, gender, pack-years of smoking, current smoking status, forced expiratory volume in one second (FEV1), CBC cell count proportions, library prep batch, and CT scanner model. Multiple comparisons were corrected with the *Benjamini-Hochberg* method to control the false discovery rate (FDR) at 10%(6). Gene ontology (GO) pathway enrichment analysis was performed using the GO “Biological Process” gene sets with p-values calculated with the Fisher exact test statistic using the weight01 algorithm in topGO (v2.33.1) (16) that accounts for dependency in GO topology(17). We reported GO pathways with at least three significant genes, and the threshold for statistical significance was an adjusted P-value < 0.001.

## Supplemental Results

As shown in **Error! Reference source not found**., both IEAs were negatively associated with emphysema peel/core distribution 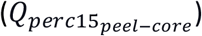, indicating that higher IEA values are associated with more emphysema in the core/central regions of the lung. To explore this association further and to determine whether this may be driven by artifacts of lung segmentation, we performed a sensitivity analysis where the lung was separated into concentric bands according to the distance to the lung boundary, as shown in Supplemental Figure E3. While the core region was always defined as the lung regions >20 mm from the lung boundary, the peel region was alternatively defined using each of the concentric bands between the core and lung periphery. For the IEA_emph_ axis, the negative correlation to 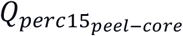 was observed for all concentric bands outside the core; however, the IEA_airway_ axis was significantly negatively correlated to peel-core emphysema only when the peel was defined as the outermost band (Supplemental Table E18). This suggests that IEA_emph_ reflects a consistently positive relationship with core emphysema, whereas, for IEA_airway_, it is not currently clear whether this reflects a real biological phenomenon at the extreme periphery of the lung, or this may be due to artifacts of segmentation.

## Supplemental Figures and Tables

**The following tables are included in the supplemental excel file:**

Table E1. Cross-validation performance in IEA training.

Table E2. Linear Regression with image-expression axes (IEAs) and COPD measurements with COPDGene visit 2 data.

Table E3. Logistic regression and Cox proportional harzard models with image-expression axes (IEAs) and COPD measurements with COPDGene visit 2 data.

Table E4. Pearson’s correlation between image-expression axes (IEAs) and COPD-related characteristics and health outcomes, measured on 1,527 subjects from another subset of the COPDGene dataset that had not been used for model training.

Table E5. Linear Regression with image-expression axes (IEAs) and COPD measurements with 1,527 subjects from another subset of the COPDGene dataset that had not been used for model training.

Table E6. Logistic regression and Cox proportional harzard models with image-expression axes (IEAs) and COPD measurements with 1,527 subjects from another subset of the COPDGene dataset that had not been used for model training.

Table E7. Characteristics of subgroups defined by dividng the Image-Expression Axes (IEAs) into quadrants.

Table E8. Covariances between Image-expression Axes (IEAs), factor analysis axes (FAs), and PCA image-only axes (PCA-Is) on COPDGene visit 1 data.

Table E9. Pearson’s correlation coefficient between image-expression axes (IEAs), factor analysis axes (FAs), PCA image only axes (PCA-Is), and health outcomes on COPDGene visit 1 data.

Table E10. Linear regression analysis with image-expression axes (IEAs) and factor analysis axes (FAs) on COPDGene visit 1 data.

Table E11. Logistic regression and Cox model with image-expression axes (IEAs) and factor analysis axes (FAs) on COPDGene visit 1 data.

Table E12. Linear regression analysis with image-expression axes (IEAs) and PCA Image Only Axes (PCA-Is) on COPDGene visit 1 data.

Table E13. Logistic regression and Cox model with image-expression axes (IEAs) and PCA Image Only Axes (PCA-Is) on COPDGene visit 1 data.

Table E14. Differentially Expressed Genes (DEGs) associated with the IEAemph (adjusted p-value <.10).

Table E15. Differentially Expressed Genes (DEGs) associated with the IEAairway (adjusted p-value <.10).

Table E16. Significant Gene Ontology enrichment terms for IEAemph (adjusted p-value < 0.001).

Table E17. Significant Gene Ontology enrichment terms for IEAairway (adjusted p-value < 0.001).

Table E18. The correlation between perc15 ratio and IEAs.

**Figure E1.**
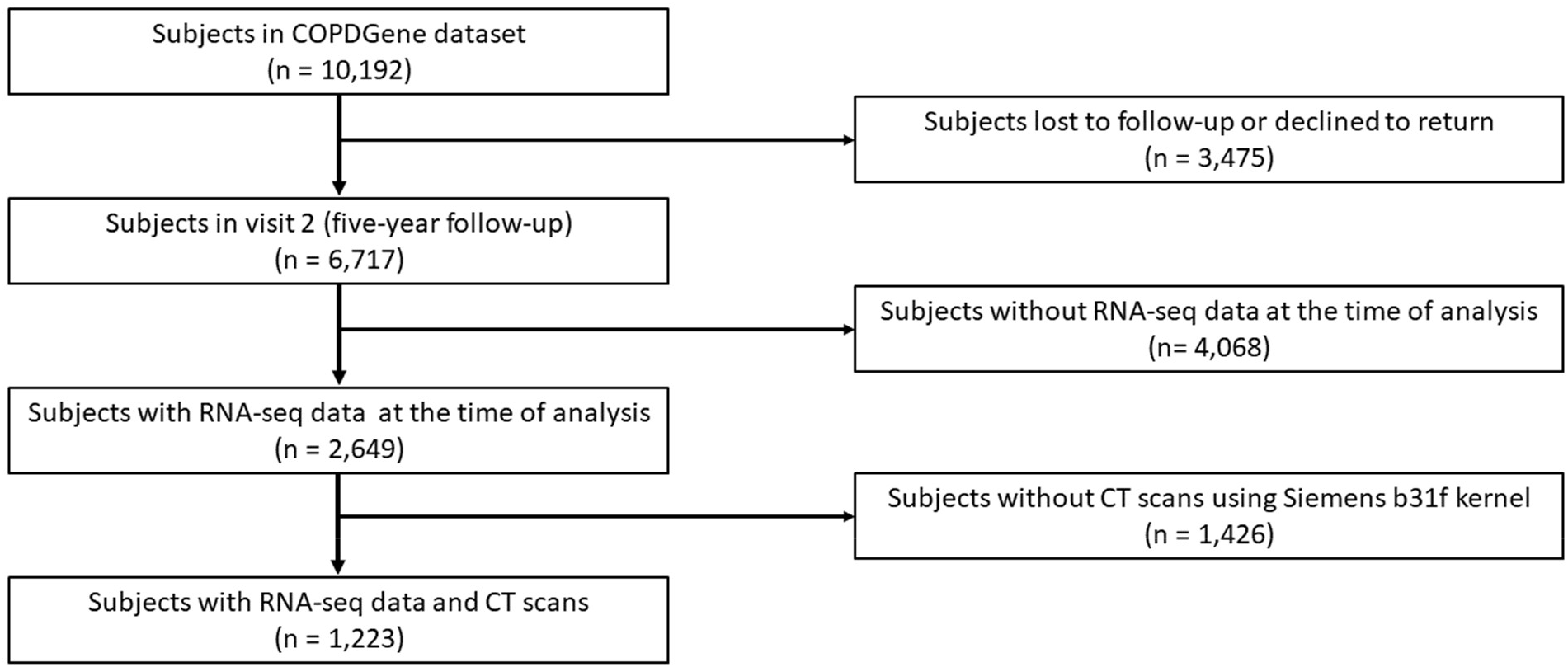
Consort Diagram showing the subjects used in the analysis.

**Figure E2.**
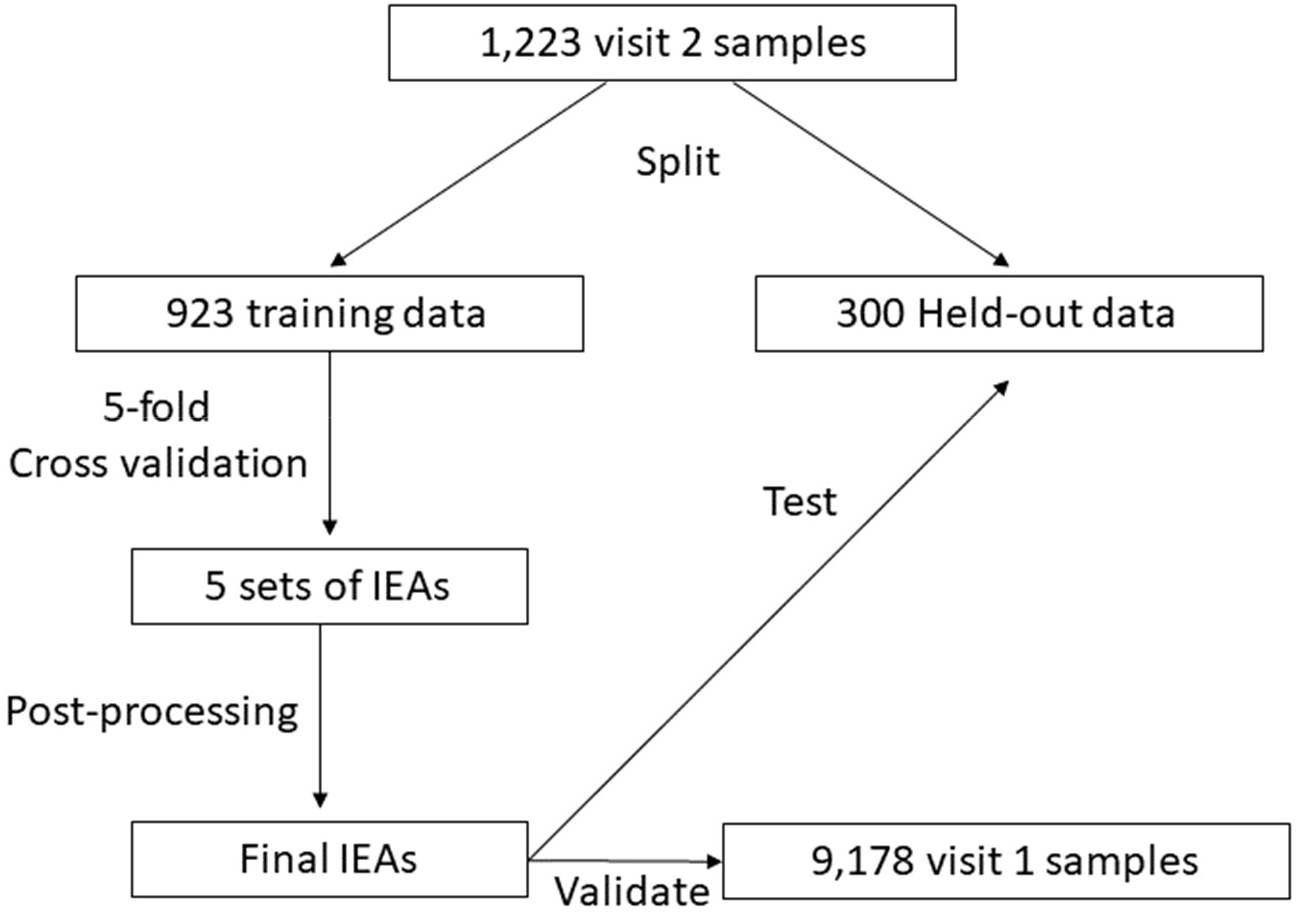
Data flow for model training and validation.

**Figure E3.**
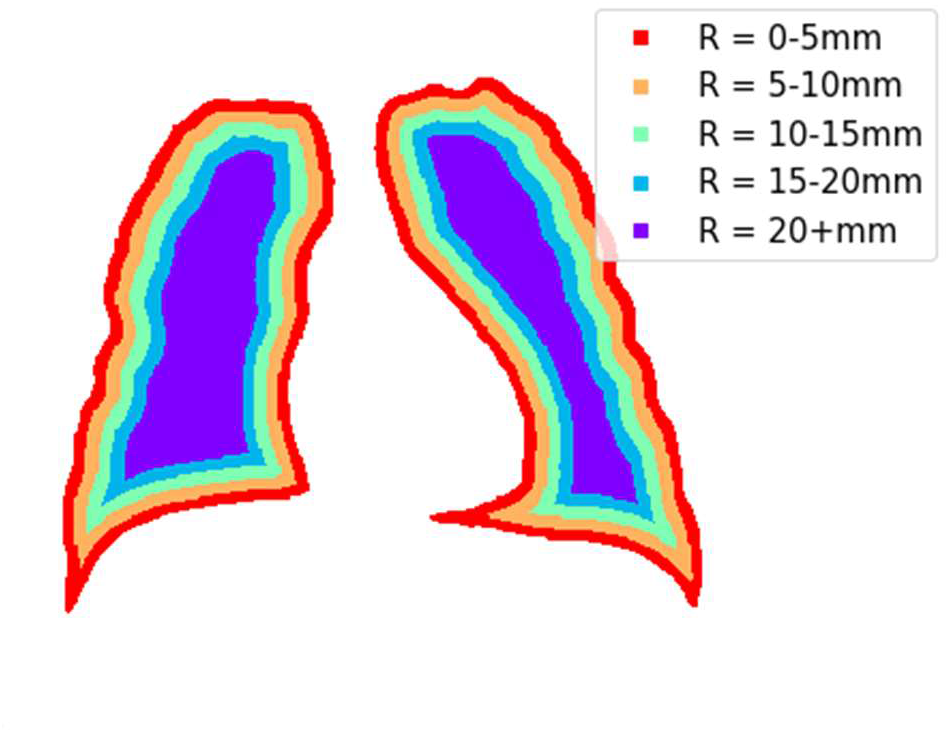
Lung “bands” used in the peel-core sensitivity analysis. To determine the robustness of the IEA association to peel-core emphysema, the 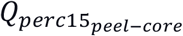 variable was recomputed using a series of lung bands defined based on the distance to the lung boundary. R is defined as the distance to the boundaries of the lung.

## References

1. Adeloye D, Chua S, Lee C, Basquill C, Papana A, Theodoratou E, et al. Global and regional estimates of COPD prevalence: Systematic review and meta–analysis. Journal of Global Health 2015;5.

2. Abubakar I, Tillmann T, Banerjee A. Global, regional, and national age-sex specific all-cause and cause-specific mortality for 240 causes of death, 1990-2013: a systematic analysis for the Global Burden of Disease Study. Lancet 2013;385:117–171.

3. Global initiative for chronic obstructive lung disease. Global Strategy for Prevention, Diagnosis, and Management of chronic obstructive pulmonary disease. 2021.

4. Rabe K, Hurd S, Anzueto A, Barnes P, Buist S, Calverley P, et al. Global Strategy for the Diagnosis, Management, and Prevention of Chronic Obstructive Pulmonary Disease. American Journal of Respiratory and Critical Care Medicine 2007;176:532–555.

5. Patel B, Coxson H, Pillai S, Agustí A, Calverley P, Donner C, et al. Airway Wall Thickening and Emphysema Show Independent Familial Aggregation in Chronic Obstructive Pulmonary Disease. American Journal of Respiratory and Critical Care Medicine 2008;178:500–505.

6. Barnes P, Celli B. Systemic manifestations and comorbidities of COPD. European Respiratory Journal 2009;33:1165–1185.

7. Sevenoaks M, Stockley R. Chronic Obstructive Pulmonary Disease, inflammation and co-morbidity – a common inflammatory phenotype? Respiratory Research 2006;7:1–9.

8. Agustí A. Systemic Effects of Chronic Obstructive Pulmonary Disease: What We Know and What We Don’t Know (but Should). Proceedings of the American Thoracic Society 2007;4:522–525.

9. Milne S, King G. Role of imaging in COPD phenotyping. Respirology 2014;20:522–523.

10. Bahr T, Hughes G, Armstrong M, Reisdorph R, Coldren C, Edwards M, et al. Peripheral Blood Mononuclear Cell Gene Expression in Chronic Obstructive Pulmonary Disease. American Journal of Respiratory Cell and Molecular Biology 2013;49:316–323.

11. Agusti A, Bel E, Thomas M, Vogelmeier C, Brusselle G, Holgate S, et al. Treatable traits: toward precision medicine of chronic airway diseases. European Respiratory Journal 2016;47:410–419.

12. Kinney GL, Santorico SA, Young KA, Cho MH, Castaldi PJ, San José Estépar R, et al. Identification of chronic obstructive pulmonary disease axes that predict all-cause mortality: the COPDGene study. American journal of epidemiology 2018;187:2109– 2116.

13. Regan E, Hokanson J, Murphy J, Make B, Lynch D, Beaty T, et al. Genetic Epidemiology of COPD (COPDGene) Study Design. COPD: Journal of Chronic Obstructive Pulmonary Disease 2011;7:32–43.

14. Sun L, Yu K, Batmanghelich K. Context matters: Graph-based self-supervised representation learning for medical images. InProceedings of the. Paper presented at: AAAI Conference on Artificial Intelligence. AAAI Conference on Artificial Intelligence 2021.

15. Hinton G. Training Products of Experts by Minimizing Contrastive Divergence. Neural Computation 2002;14:1771–1800.

16. Gretton A, Bousquet O, Smola A, Schölkopf B. Measuring statistical dependence with Hilbert-Schmidt norms. Paper presented at: International conference on algorithmic learning theory 2005; Berlin, Heidelberg.

17. Kingma D, Ba J. Adam: A method for stochastic optimization. 2014. arXiv:1412.6980.

18. Cox D. Regression Models and Life-Tables. Journal of the Royal Statistical Society: Series B (Methodological) 1972;34:187–202.

19. Young KA, Regan EA, Han MK, Lutz SM, Ragland M, Castaldi PJ, et al. Subtypes of COPD have unique distributions and differential risk of mortality. Chronic Obstructive Pulmonary Diseases: Journal of the COPD Foundation 2019;6:400.

20. Ritchie M, Phipson B, Wu D, Hu Y, Law C, Shi W, et al. limma powers differential expression analyses for RNA-sequencing and microarray studies. Nucleic Acids Research 2015;43:e47–e47.

21. Phipson B, Lee S, Majewski I, Alexander W, Smyth G. Robust hyperparameter estimation protects against hypervariable genes and improves power to detect differential expression. The Annals of Applied Statistics 2016;10:946.

22. Benjamini Y, Hochberg Y. Controlling the False Discovery Rate: A Practical and Powerful Approach to Multiple Testing. Journal of the Royal Statistical Society: Series B (Methodological) 1995;57:289–300.

23. Alexa A, Rahnenführer J. Gene set enrichment analysis with topGO. Bioconductor Improv 2009;27:1–26.

24. Castaldi PJ, Cho MH, San José Estépar R, McDonald M-LN, Laird N, Beaty TH, et al. Genome-wide association identifies regulatory Loci associated with distinct local histogram emphysema patterns. American journal of respiratory and critical care medicine 2014;190:399–409.

25. McCarthy C, Reeves EP, McElvaney NG. The role of neutrophils in alpha-1 antitrypsin deficiency. Annals of the American Thoracic Society 2016;13:S297–S304.

26. Faner R, Cruz T, Casserras T, López-Giraldo A, Noell G, Coca I, et al. Network analysis of lung transcriptomics reveals a distinct B-cell signature in emphysema. American journal of respiratory and critical care medicine 2016;193:1242–1253.

27. Papaioannou A, Mazioti A, Kiropoulos T, Tsilioni I, Koutsokera A, Tanou K, et al. Systemic and airway inflammation and the presence of emphysema in patients with COPD. Respiratory Medicine 2010;104:275–282.

28. Tkacova R. Systemic Inflammation in Chronic Obstructive Pulmonary Disease: May Adipose Tissue Play a Role? Review of the Literature and Future Perspectives. Mediators of Inflammation 2010;2010:1–11.

29. Mair G, Miller JJ, McAllister D, Maclay J, Connell M, Murchison JT, et al. Computed tomographic emphysema distribution: relationship to clinical features in a cohort of smokers. European Respiratory Journal 2009;33:536–542.

30. Gurney JW, Jones KK, Robbins RA, Gossman GL, Nelson KJ, Daughton D, et al. Regional distribution of emphysema: correlation of high-resolution CT with pulmonary function tests in unselected smokers. Radiology 1992;183:457–463.

31. Nakano Y, Sakai H, Muro S, Hirai T, Oku Y, Nishimura K, et al. Comparison of low attenuation areas on computed tomographic scans between inner and outer segments of the lung in patients with chronic obstructive pulmonary disease: incidence and contribution to lung function. Thorax 1999;54:384–389.

32. Haraguchi M, Shimura S, Hida W, Shirato K. Pulmonary function and regional distribution of emphysema as determined by high-resolution computed tomography. Respiration 1998;65:125–129.

33. Boueiz A, Chang Y, Cho MH, Washko GR, Estépar RSJ, Bowler RP, et al. Lobar emphysema distribution is associated with 5-year radiological disease progression. Chest 2018;153:65–76.

34. Yang J, Angelini E, Balte P, Hoffman E, Austin J, Smith B, et al. Novel Subtypes of Pulmonary Emphysema Based on Spatially-Informed Lung Texture Learning: The Multi-Ethnic Study of Atherosclerosis (MESA) COPD Study. IEEE Transactions on Medical Imaging 2021;40:3652–3662.

35. Ohri K, Kumar M. Review on self-supervised image recognition using deep neural networks. Knowledge-Based Systems 2021;224:107090.

36. Cho M, Castaldi P, Hersh C, Hobbs B, Barr R, Tal-Singer R, et al. A genome-wide association study of emphysema and airway quantitative imaging phenotypes. American journal of respiratory and critical care medicine 2015;192(5):559–569.

37. Jeong I, Lim J-H, Oh DK, Kim WJ, Oh Y-M. Gene expression profile of human lung in a relatively early stage of COPD with emphysema. International Journal of Chronic Obstructive Pulmonary Disease 2018;13:2643.

## References

1. Regan E, Hokanson J, Murphy J, Make B, Lynch D, Beaty T, et al. Genetic Epidemiology of COPD (COPDGene) Study Design. COPD: Journal of Chronic Obstructive Pulmonary Disease 2011;7:32–43.

2. Tustison NJ, Cook PA, Klein A, Song G, Das SR, Duda JT, et al. Large-scale evaluation of ANTs and FreeSurfer cortical thickness measurements. Neuroimage 2014;99:166–179.

3. Sun L, Yu K, Batmanghelich K. Context matters: Graph-based self-supervised representation learning for medical images. InProceedings of the. Paper presented at: AAAI Conference on Artificial Intelligence. AAAI Conference on Artificial Intelligence 2021.

4. Li Y, Van De Geijn B, Raj A, Knowles D, Petti A, Golan D, et al. RNA splicing is a primary link between genetic variation and disease. Science 2016;352:600–604.

5. Robinson M, Mccarthy D, Smyth G. edgeR: a Bioconductor package for differential expression analysis of digital gene expression data. Bioinformatics 2010;26:139–140.

6. Benjamini Y, Hochberg Y. Controlling the False Discovery Rate: A Practical and Powerful Approach to Multiple Testing. Journal of the Royal Statistical Society: Series B (Methodological) 1995;57:289–300.

7. Hinton G. Training Products of Experts by Minimizing Contrastive Divergence. Neural Computation 2002;14:1771–1800.

8. Gretton A, Bousquet O, Smola A, Schölkopf B. Measuring statistical dependence with Hilbert-Schmidt norms. Paper presented at: International conference on algorithmic learning theory 2005; Berlin, Heidelberg.

9. Kingma D, Ba J Adam. Adam: A method for stochastic optimization. 2014.

10. Kingma DP, Welling M. Auto-encoding variational bayes. 2013. arXiv preprint arXiv:1312.6114

11. Cox D. Regression Models and Life-Tables. Journal of the Royal Statistical Society: Series B (Methodological) 1972;34:187–202.

12. Castaldi PJ, Cho MH, San José Estépar R, McDonald M-LN, Laird N, Beaty TH, et al. Genome-wide association identifies regulatory Loci associated with distinct local histogram emphysema patterns. American journal of respiratory and critical care medicine 2014;190:399–409.

13. Young KA, Regan EA, Han MK, Lutz SM, Ragland M, Castaldi PJ, et al. Subtypes of COPD have unique distributions and differential risk of mortality. Chronic Obstructive Pulmonary Diseases: Journal of the COPD Foundation 2019;6:400.

14. Ritchie M, Phipson B, Wu D, Hu Y, Law C, Shi W, et al. limma powers differential expression analyses for RNA-sequencing and microarray studies. Nucleic Acids Research 2015;43:e47–e47.

15. Phipson B, Lee S, Majewski I, Alexander W, Smyth G. Robust hyperparameter estimation protects against hypervariable genes and improves power to detect differential expression. The Annals of Applied Statistics 2016;10:946.

16. Alexa A, Rahnenführer J. Gene set enrichment analysis with topGO. Bioconductor Improv 2009;27:1–26.

17. Alexa A, Rahnenfuhrer J, Lengauer T. Improved scoring of functional groups from gene expression data by decorrelating GO graph structure. Bioinformatics 2006;22:1600–1607.

